# Predicting the outcomes of Kasai portoenterostomy for biliary atresia: a cohort study

**DOI:** 10.1101/2022.10.06.22279593

**Authors:** Qiao Qi, Yanfu Wang, Qijun Wu, Pengjun Su, Dajia Wang, Tianyu Li, Zhibo Zhang

**Affiliations:** Department of Pediatric Surgery, Shengjing Hospital of China Medical University; Department of Epidemiology, Shengjing Hospital of China Medical University

## Abstract

**Objectives:** To identify factors associated with outcomes of Kasai portoenterostomy (KPE), and predictors of 2- and 5- year native liver survival (NLS) for infants achieved jaundice clearance (JC) within 6 months of KPE.

**Methods:** This retrospective cohort study was conducted on 151 patients with type III biliary atresia (BA) who underwent KPE at our center. Univariate analysis and logistic regression analyses were performed to identify factors associated with NLS in infants achieved JC. Kaplan–Meier curves and log-rank tests were used to estimate the NLS, and the Cox proportional hazards regression model identified variables most associated with 2- and 5-year NLS at 6 months post-KPE. A receiver operating characteristic (ROC) curve was used to evaluate the predictive value of these factors.

**Results:** The 2- and 5-year NLS of infants achieved JC at 3 months post-KPE were not different from those achieved JC earlier. Operation age and total bile acid (TBA) were factors associated with JC. For infants who have achieved JC, DB was the only factor associated with 2-year NLS, the AUC was 0.872, the cutoff value was 14 μmol/L; ALB and DB were factors associated with 5-year NLS, the AUCs were 0.894 and 0.95, and the cutoff values were 39 g/L and 14 μmol/L, respectively.

**Conclusions:** NLS should be estimated at 6 months post-KPE. Preoperative factors are not predictive of NLS. For infants cleared jaundice, DB and ALB can predict NLS with good performance.

**What’s Known on This Subject:** Age, liver stiffness, and CMV infections are factors associated with outcomes of Kasai portoenterostomy. Jaundice clearance is directly associated with native liver survival; however, even with successful surgery, liver pathology in most cases will progress to end-stage cirrhosis.

**What This Study Adds:** No preoperative factors are predictive of native liver survival (NLS). Infants cleared jaundice after 3 months of KPE can achieve the same NLS as those cleared jaundice earlier. For infants cleared jaundice, 6-month postoperative DB and Albumin are predictive of NLS.

**How this study might affect research, practice or policy:** In this study, we argued that 6 months post-KPE was the appropriate timing for predicting NLS; direct bilirubin (DB) and albumin (ALB) at 6 months post-KPE can be used to predict 2- and 5-year NLS with good performance.

**Article Summary:** Retrospective analysis revealed it’s difficult to predict outcomes of Kasai portoenterostomy (KPE) preoperatively; jaundice clearance should be evaluated at 6 months after KPE, for infants cleared jaundice, 6-month postoperative DB and Albumin are predictive of NLS.

## Introduction

Biliary atresia (BA) is a devastating neonatal cholangiopathy.^1^ Affected infants suffer from persistent and progressive cholestatic jaundice beyond the neonatal period, which can lead to life-threatening end-stage liver failure if left untreated. The incidence of BA varies across different areas aroud the world.^2^ Kasai portoenterostomy (KPE) is the primary surgical treatment for BA, although primary liver transplantation (LT) has been advocated by some surgeons. Successful KPE can re-establish biliary drainage and achieve long-term native liver survival (NLS). However, only approximately 40–70% of infants achieve jaundice clearance (JC) after KPE, and those who fail to achieve JC will require LT shortly afterwards. Even in the case of jaundice resolution, liver fibrosis progresses in most infants, who eventually require transplantation at a later age.^3^ BA is currently the most common cause of pediatric LTs.^4^

Most studies analyzing the prognosis of BA engaged in prediction of JC and NLS using preoperative and immediate postoperative factors, but few studies have been reported about the predictors for patients who have achieved JC. By 6 months following KPE, infants who do not have optimal bilirubin are commonly predicted to require LT. However, for infants who have achieved JC, how to predict their probability of NLS is a pressing question for surgeons. In this study, we aimed to analyze the possible factors associated with KPE outcomes and to identify prognostic factors that may be used to predict NLS in infants who have achieved JC. We hypothesized that combined clinical and laboratory measures reflecting the overall conditions of the liver can enhance the ability to predict NLS.

## Methods

### Research subjects

This single-center retrospective cohort study was performed at a tertiary pediatric center in Northeast China. The medical records of all infants who underwent KPE between January 2011 and June 2021 for type III BA were collected. Ethical approval for this study was obtained from the Research Ethics Committee of the Hospital (2021PS008K).

Patients were diagnosed using intraoperative cholangiography and/or liver pathology. As reported by Nio et al. in 2016,^5^ fibrous remnants were transected at the hepatic capsule level, and the Roux-en Y procedure with a spur valve was employed for all patients. The length of the Roux-en Y loop ranged from 25 to 30 cm, according to the distance from the umbilicus to the hilum. Postoperative adjuvant therapy included corticosteroids, antibiotics to prevent cholangitis, ursodeoxycholic acid, nutritional support, and supplementation of fat-soluble vitamins. Follow-up was maintained until LT, death, or cessation of visits, whichever occurred first. Data collection was completed in January 2022.

### Data collection and outcome definition

Data, including sex, age at KPE, pre- and post-operative laboratory examinations (alanine aminotransferase [ALT], aspartate aminotransferase [AST], gamma-glutamyl transpeptidase [GGT], albumin [ALB], total bilirubin [TB], direct bilirubin [DB], total bile acid [TBA], and platelet [PLT]), virology examination results, liver elasticity, and liver fibrosis at pathology, were collected.

JC was defined as TB<20 μmol/L within 6 months of KPE, and infants were divided into jaundice clearance (JC) and jaundice unclearance (JUC) groups. Infants in the JC group were further divided into early jaundice clearance (EJC) group (cleared jaundice within 3 months) and late jaundice clearance (LJC) group (cleared jaundice between 3 and 6 months), based on the assumption that the sooner jaundice is cleared, the better the prognosis.

Patients who achieved JC within 6 months post-KPE were divided into 2-year NLS and 2-year non NLS groups, and 5-year NLS and 5-year non NLS groups, according to the time post-KPE as well as survival with native liver or LT/death.

### Statistical analysis

The study was powered to detect differences in clinical parameters between groups. Univariate analysis of clinical and laboratory data was performed using the chi-square and Student’s *t*-tests. Variables with *P*<0.05 were included in a multiple logistic regression model to select the best variables associated with JC.

Kaplan–Meier curves and log-rank tests were used to estimate the 2- and 5-year NLS, and variables with *P*<0.1 were included in the Cox proportional hazards regression model to select predictive variables for 2- and 5-year NLS. Factors reflecting different liver functions were also included in the model.

Receiver operating characteristic (ROC) analysis of variables in the regression model was performed to determine optimal cut-off values (to maximize both sensitivity and specificity) for predicting 2- and 5-year NLS. The performance of the regression model was assessed using the area under the ROC curve (AUC), and the results are expressed as hazard ratios (HRs) with 95% confidence intervals (CIs).

Statistical analyses were performed using SPSS version 23.0 (IBM Corp., Armonk, NY, USA), MedCalc version 12.7.0 (MedCalc Software, Ostend, Belgium), and GraphPad Prism version 8.0 (GraphPad Software, San Diego, CA, USA). Statistical significance was set at *P*<0.05.

## Results

### General clinical data

General clinical data was listed in table 1.

**Table 1.**
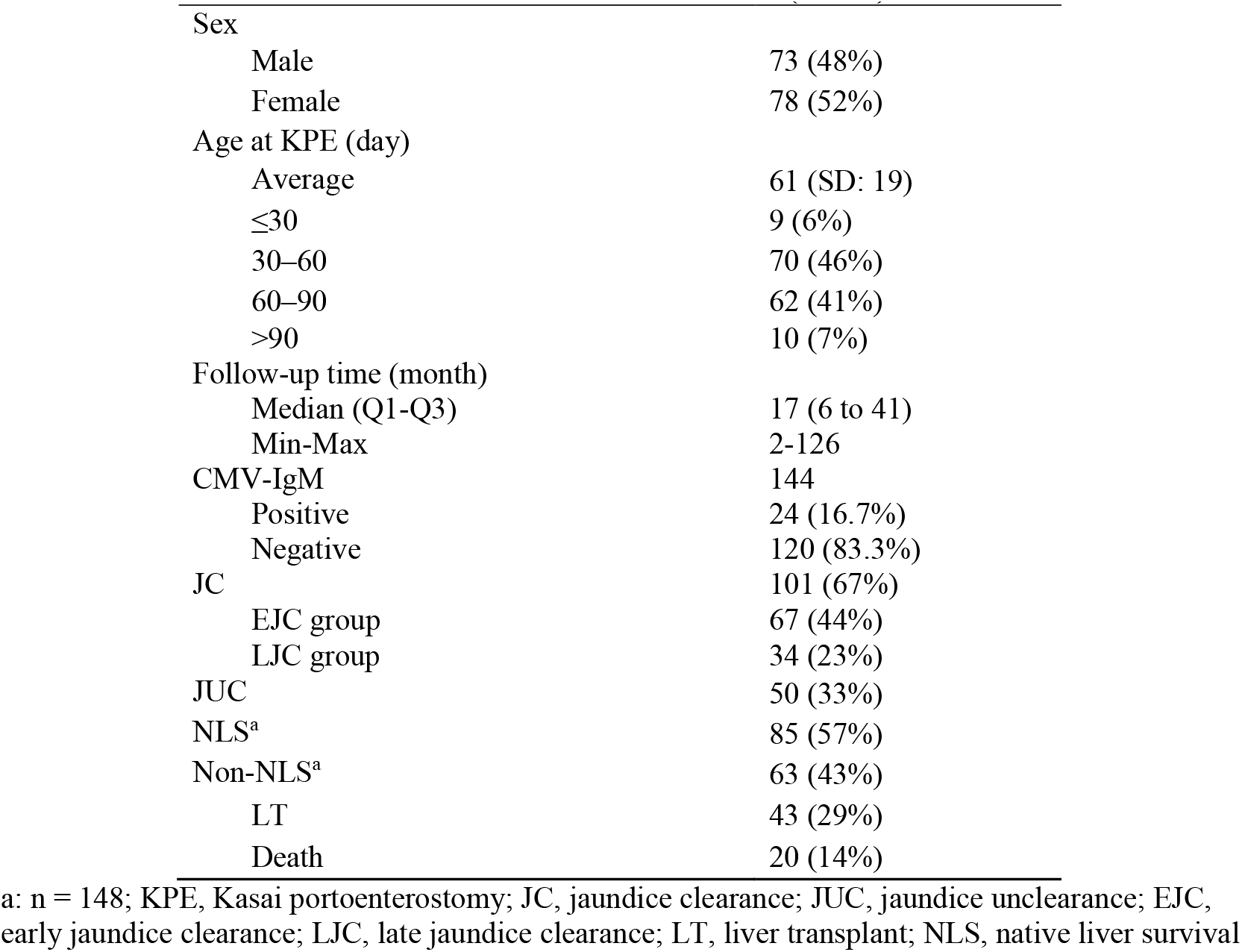
General clinical data (n=151)

|  |  |
| --- | --- |
| Sex |  |
| Male | 73 (48%) |
| Female | 78 (52%) |
| Age at KPE (day) |  |
| Average | 61 (SD: 19) |
| ≤30 | 9 (6%) |
| 30–60 | 70 (46%) |
| 60–90 | 62 (41%) |
| >90 | 10 (7%) |
| Follow-up time (month) |  |
| Median (Q1-Q3) | 17 (6 to 41) |
| Min-Max | 2-126 |
| CMV-IgM | 144 |
| Positive | 24 (16.7%) |
| Negative | 120 (83.3%) |
| JC | 101 (67%) |
| EJC group | 67 (44%) |
| LJC group | 34 (23%) |
| JUC | 50 (33%) |
| NLS<sup>a</sup> | 85 (57%) |
| Non-NLS<sup>a</sup> | 63 (43%) |
| LT | 43 (29%) |
| Death | 20 (14%) |

a: n = 148; KPE, Kasai portoenterostomy; JC, jaundice clearance; JUC, jaundice unclearance; EJC, early jaundice clearance; LJC, late jaundice clearance; LT, liver transplant; NLS, native liver survival

Overall, 151 patients were enrolled in this study. JC was achieved in 101 infants (66.9%) within 6 months of KPE. Three cases that achieved JC were excluded from the analysis of 2- and 5-year NLS because they were lost to follow-up between the 6- and 1-year post-KPE. By the end of the follow-up period, 85 infants survived with their native liver (57.4%), 43 underwent LT, and 20 died without LT.

### Factors associated with JC

Univariate analysis was performed to compare clinical data between the JC (n = 101) and JUC (n = 50) groups. Results showed that age at KPE, AST, TBA, and APRI were associated with JC; JCs was the highest in the <30 d group (100%) followed by the 30–60 d (74%), 60–90 d (60%), and >90 d (30%) groups, while the difference between the <30 d and 30–60 d groups was not significant. We compared JC between ≤60 d group and >60 d group and found that the JC rate in ≤60 d group was higher than that in >60 d group (Table 2).

**Table 2.**
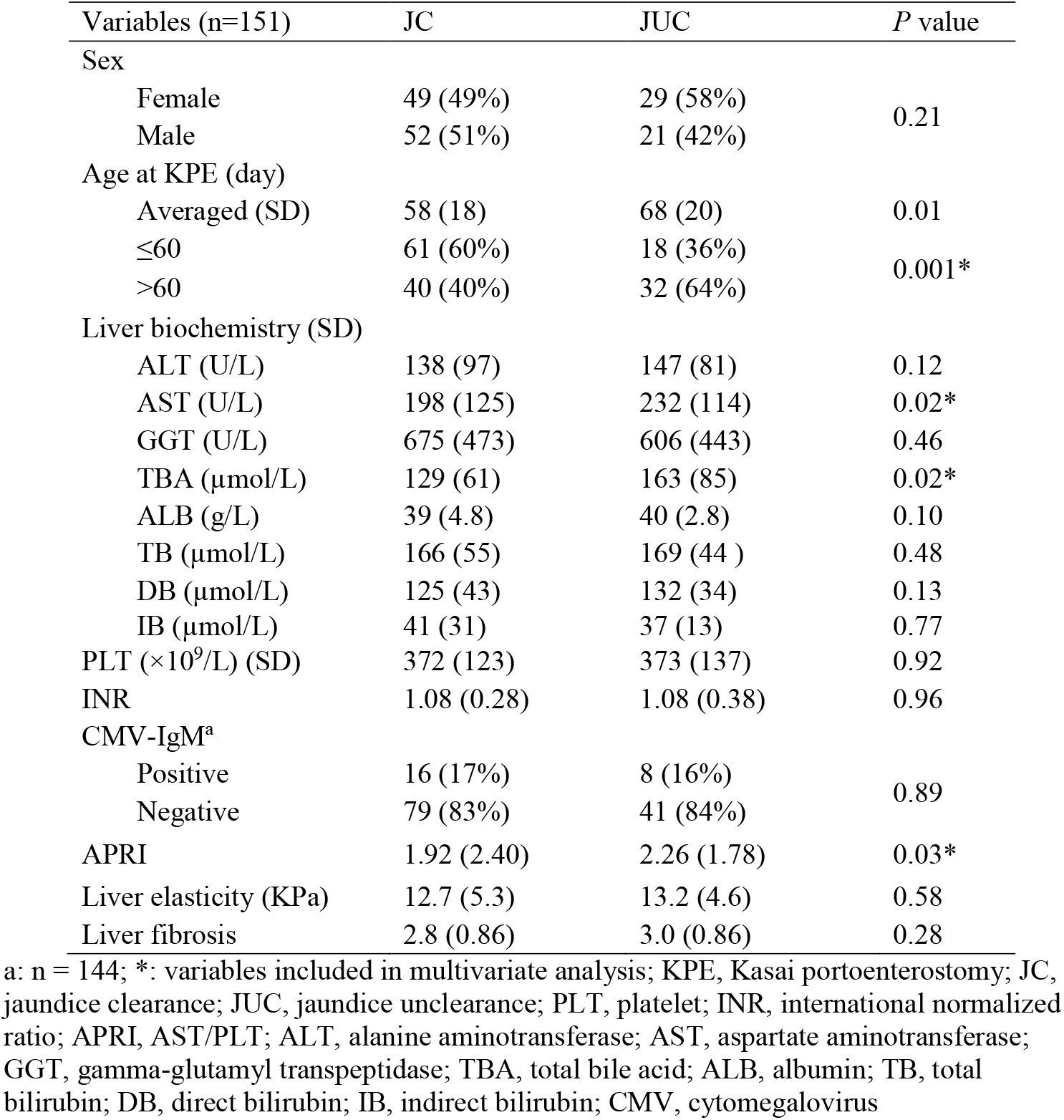
Univariate analysis of the association between jaundice clearance and subject demographic and clinical factors.

In the multivariate logistic analysis, only age at KPE and TBA were associated with JC, although both of these were independent risk factors for JC (OR 1.021, 95% CI 1.001–1.042, *P*=0.04; OR 1.005, 95% CI 1.000–1.011, *P*=0.04), the AURs were no more than 0.7 (online supplemental Fig S1).

### Factors associated with NLS for the total cohort

113 children were included in the 2-year NLS analysis cohort, of which 54.9% (62 cases, including 6 who did not achieve complete JC) survived with native liver and the other 45.1% (51 cases) either underwent LT or died. 62 children were included in the 5-year NLS analysis cohort, of which 41.9% (26 cases, including 1 case who did not achieve complete JC) survived with native liver and the other 58.1% (36 cases) underwent LT or died. Kaplan–Meier curves showed that most deaths/LT events occurred within 1 year of KPE (online supplemental Fig S2).

Univariate analysis revealed that there was no difference in the ages at KPE between the NLS and non-NLS groups (60 d vs. 67 d, *P*=0.09); however, 2-year NLS was different in various age groups. The highest rate was in the <30 d group (75%), followed by the 30–60 d (63%), 60–90 d (52%), and the >90 d (13%) groups (*P*=0.005) (Fig 1). Another factor associated with 2-year NLS was JC (*P*=0.000).

**Figure 1.**
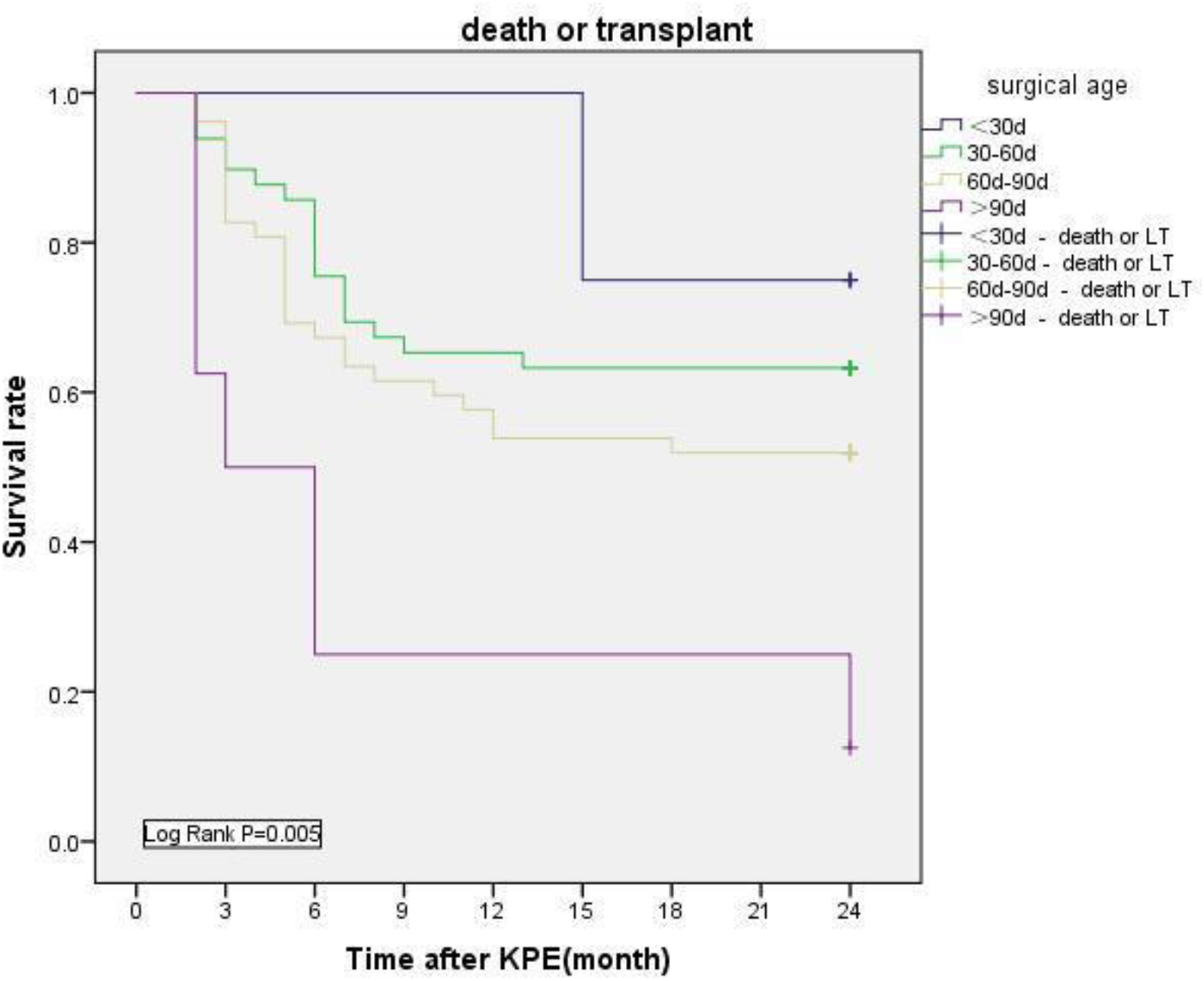
Kaplan–Meier survival curve and log-rank analysis of age at surgery on 2-year NLS

Univariate analysis of the 5-year NLS revealed that sex (males 48% vs. females 52%, *P*=0.007) and JC (JC 60% vs. JUC 40%, *P*=0.000) were the only factors associated with the 5-year NLS.

The Cox proportional hazards regression model was used to analyze possible risk and protective factors for 2- and 5-year NLS, and sex, age at KPE, and JC were included according to the results of univariate analysis. The results showed that uncleared jaundice was the only risk factor for both 2- and 5-year NLS (HR 8.792, 95% CI 4.576–16.893, *P*=0.000; HR 9.316, 95% CI 4.143–20.95, *P*=0.000).

### Predicting NLS for patients who have achieved JC

Among infants achieved JC, 68 infants underwent surgery for more than two years and 36 infants for more than five years were enrolled. According to whether they lived with native liver or underwent LT/died, they were further divided into the 2-year NLS group (n=56) and the 2-year non-NLS group (n=12), 5-year NLS group (n=25) and the 5-year non-NLS group (n=11). Infants who failed to achieve complete JC but survived with native liver for 2 (n=6) and 5 years (n=1) are not included in the analysis.

The routine laboratory and clinical factors at 6 months post-KPE, which included TB, DB, TBA (indicating the bile excretory function of the liver), ALT (indicating hepatocyte destruction), ALB (indicating the albumin synthesis ability of the liver), and APRI (indicating the degree of liver fibrosis), were analyzed between groups.

In univariate analysis, sex, ALB, TB, and DB were associated with 2-year NLS (Table 3); after analysis with Cox hazard regression model, DB was the only factor independently associated with 2-year NLS (HR 1.111, 95% CI 1.033–1.193, *P*=0.004). ROC analysis revealed an optimal cutoff value for DB <14 µmol/L (AUC 0.872, Youden index 0.67, P<0.001) to predict 2-year NLS (online supplemental Table S1, Fig S3A).

**Table 3.** Univariate analysis of factors associated with 2- and 5-year NLS in JC infants.

|  | 2-year (n = 68) |  |  | 5-year (n = 36) |  |  |
| --- | --- | --- | --- | --- | --- | --- |
|  | NLS group | Non-NLS group | *P* value | NLS group | Non-NLS group | *P* value |
| Sex |  |  |  |  |  |  |
| Male | 34 (61%) | 3 (25%) | 0.03* | 18 (72%) | 2 (18%) | 0.007* |
| Female | 22 (39%) | 9 (75%) |  | 7 (28%) | 9 (82%) |  |
| Age at KPE (day) (SD) | 59 (16) | 61 (21) | 0.66 | 61 (14) | 67 (17) | 0.27 |
| Rapidity of JC |  |  |  |  |  |  |
| EJC | 47 | 11 | 0.501 | 22 | 11 | 0.999 |
| LJC | 9 | 1 |  | 3 | 0 |  |
| Liver function biochemistry (SD) |  |  |  |  |  |  |
| ALT (U/L) | 120 (106) | 222 (236) | 0.07 | 106 (112) | 193 (156) | 0.12 |
| ALB (g/L) | 41 (4.3) | 37 (2.8) | 0.03* | 42 (4.5) | 37 (1.7) | 0.02* |
| TB (μmol/L) | 11.6 (10.4) | 25.4 (10.0) | 0.003* | 10.0 (8.0) | 31.5 (12.4) | 0.004* |
| DB (μmol/L) | 7.8 (8.9) | 20.5 (9.4) | 0.002* | 6.0 (5.6) | 26.4 (11.0) | 0.006* |
| TBA (μmol/L) | 61 (127) | 71 (41) | 0.83 | 27 (30) | 69 (39) | 0.02* |
| APRI (SD) | 1.74 (1.58) | 4.30 (6.59) | 0.11 | 1.536 (1.77) | 1.52 (SD: 1.30) | 0.98 |

\*: factors included in multivariable analysis.
JC, jaundice clearance; EJC, early jaundice clearance; LJC, late jaundice clearance; NLS, native liver survival; KPE, Kasai portoenterostomy; ALT, alanine aminotransferase; ALB, albumin; TB, total bilirubin; DB, direct bilirubin; TBA, total bile acid; APRI, AST/PLT

Univariate analysis showed that sex, TBA, ALB, TB, and DB were factors associated with 5-year NLS (Table 3). After analysis with the Cox proportional hazards regression model, ALB and DB were found to be associated with 5-year NLS (HR 0.705, 95% CI 0.526–0.944, *P*=0.02; HR 1.058, CI 1.004– 1.116, *P*=0.04). ROC analysis revealed an optimal cutoff value of Alb >39 g/L (AUC 0.894, Youden index 0.85, *P*=0.001) and DB <14 µmol/L (AUC 0.95, Youden index 0.839, *P*<0.001) for prediction of 5-year NLS (online supplemental Table S1, Fig S3B).

### Infants of LJC group can achieve the same NLS as those of EJC group

To prove infants achieved JC beyond 3 months of KPE can achieve the same prognosis with those who achieved JC earlier, we compared the 2- and 5-year NLS between EJC and LJC group, and found that the 2- and 5-year NLS of the two groups were not different (Table 3).

## Discussion

KPE remains the first surgical choice for biliary atresia, considerable efforts have been expended to optimize the results of hepatoportoenterostomy.^3-8^ However, not all efforts have proved to be efficacious, and most patients eventually require LT or even die.

JC and NLS are two main outcomes of KPE for infants with BA. Infants who do not achieve JC are commonly at a high risk for LT; for those who cleared jaundice, how to predict long-term survival remains unclear, which is also a pressing question for surgeons. In this study, we found that no preoperative factors are predictive of 2-year and 5-year NLS for infants achieved JC, however, DB, DB combined with ALB 6-months post-KPE can predict 2- and 5- year NLS with good performance. DB and ALB are factors reflecting the excretory and synthetic functions of liver; the results indicate that only patients with excellent comprehensive liver functions can survive with their native liver. Some studies have been reported about predicting NLS for infants after KPE, most focused on factors before jaundice resolution, and some are not routine parameters,^9-15^ the predictive values are not certain. Studies have reported that a rapid decrease in TB levels within 3 months of KPE accurately predicts NLS in BA, ^10^ however, we did not discover a correlation between rapid decline of bilirubin and NLS, infants who cleared jaundice beyond 3 months showed the same NLS as those achieved JC earlier. Therefore, we argue that it is not appropriate to evaluate the NLS at 3 months post-KPE or earlier. We also argue that DB, instead of TB, and ALB were predictors for 2- and 5-year NLS, which indicates that only children with optimal excretory, synthetic, and conjugating functions can achieve long-term NLS.

Undoubtedly, restoration of adequate bile flow and achievement of complete JC was the primary result to be concerned.^2, 16-19^ In the present study, we found that JC was associated with younger operative age, and infants who underwent KPE within 60 days of birth had better JC than that of infants older than 60 days; therefore, we propose that the optimal window for KPE is within 60 days.

The ultimate goal of KPE is to achieve long-term NLS and postpone the need for LT.^20^ Reported NLS ranges from 30% to 88% worldwide.^21-25^ In the present study, the 2- and 5- year NLS rates in our patients were 54.9% and 41.9%, respectively, we can see that NLS declined mainly in the first year. Failed KPE was the main reason for the decline, and the other important reason was cholangitis, especially in the early days of the study. Currently, LT due to cholangitis is becoming increasingly rare. Although the impact of cholangitis on NLS was great, we did not discuss the topic in this paper, because the incidence of this cohort was high and the treatment strategy was consistent.

When analyze the NLS of the total cohort, we found that only JC and sex were associated with NLS, which indicates that only infants with optimal bilirubin levels are likely to achieve NLS; this is why prediction of NLS in JC infants is imperative. NLS in this study was higher in boys than in girls, the reason is unclear.

Many studies have reported that age at KPE is a major factor correlated with NLS. Li et al. reported that each additional day of age toward KPE was associated with a 2% decrease in the likelihood of NLS.^25^ Consistent with other reports,^24-27^ we found that the age was associated with higher ORs for JC; however, we did not reveal the impact of age at KPE on the 2- and 5-year NLS, which may be due to the fact that few patients older than 90 days were included in this study. Additionally, we did not find associations between CMV-IgM and JC or NLS, which was inconsistent with reports by others.^28, 29^

The present study has several limitations. Given its retrospective nature, incomplete data limited the number of patients that could be used in the analysis of each variable. Another limitation was possible data bias due to this being a single center study.

The strength of this study was the consistency and continuity of the cohort; consequently, the rate of lost-to-follow-up was very low. As a single-center study, all patients were treated following the same principle and perioperative management. Therefore, the background of the study was consistent across a long period of time.

Long-term complications are common in BA patients with native liver, and timely and effective management of these complications cannot be overstated. Optimal NLS are always reported by centers with meticulous perioperative care and close follow-up.^24, 30-31^

In conclusion, predicting the outcomes of KPE is pressing and challenging for surgeons treating BA. Age at KPE, preoperative TBA and APRI are factors associated with JC; however, except for JC, none of the preoperative parameters were effective in predicting NLS. Prediction of NLS was more significant in infants who have achieved JC; the DB and ALB at 6 months after KPE are significant predictors of long-term NLS.

## Data Availability

All data produced in the present study are available upon reasonable request to the authors
All data produced in the present work are contained in the manuscript

## Abbreviations

KPE: Kasai portoenterostomy
BA: biliary atresia
JC: jaundice clearance
JUC: jaundice unclearance
LT: liver transplantation
NLS: native liver survival
PLT: platelet
INR: international normalized ratio
ALT: alanine aminotransferase
AST: aspartate aminotransferase
GGT: gamma-glutamyl transpeptidase
TBA: total bile acid
ALB: albumin
TB: total bilirubin
DB: direct bilirubin
IB: indirect bilirubin
APRI: AST/PLT
CMV: cytomegalovirus
ROC: receiver operating characteristic curve
AUC: area under the ROC

