## supplemental table S1, Figure S1-S3 for "Predicting the outcomes of Kasai portoenterostomy for biliary atresia: a cohort study"

**Table S1. Predicting performance of ALB and DB on 2- and 5-year NLS of JC patients**

|  | Variable | AUC | PPV (%) | NPV (%) | Sensitivity | Specificity | Youden index | Cut-off value | *P* value |
| --- | --- | --- | --- | --- | --- | --- | --- | --- | --- |
| For 2-year NLS | DB | 0.872 | 0.571 | 0.952 | 0.8 | 0.87 | 0.67 | 14 | ＜0.001 |
| For 5-year NLS | ALB | 0.894 | 0.75 | 1 | 1 | 0.15 | 0.85 | 39 | 0.001 |
|  | DB | 0.95 | 0.889 | 0.95 | 0.889 | 0.95 | 0.839 | 14 | <0.001 |

JC, jaundice clearance; DB, direct bilirubin; NLS, native liver survival; AUC: area under the receiver characteristic curve; PPV: positive predictive value; NPV: negative predictive value

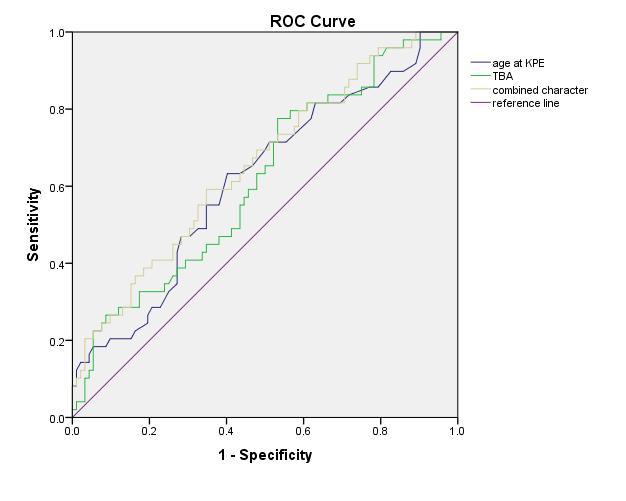

Figure S1. ROC curve of variables associated with JC

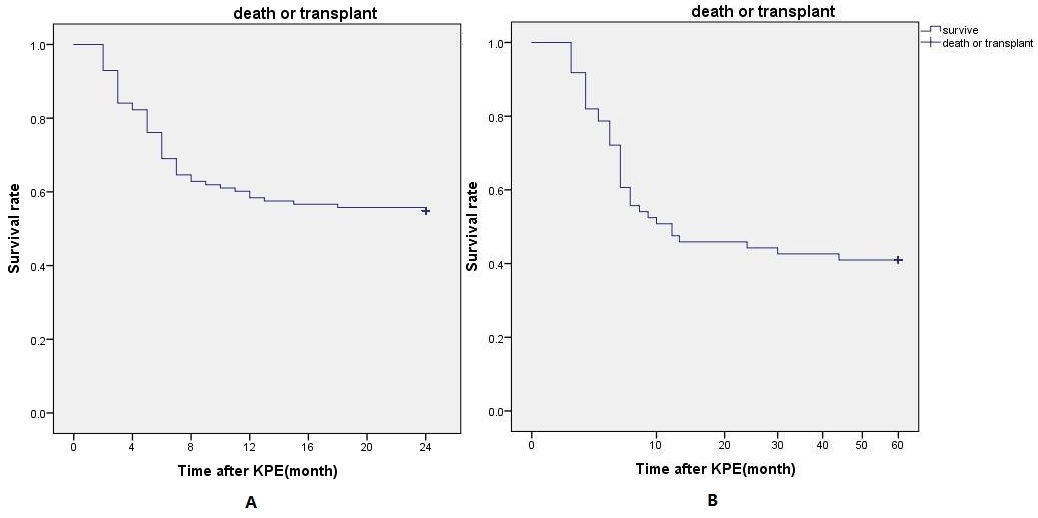

Figure S2. Kaplan–Meier survival curve of 2- (A) and 5-year NLS (B) after KPE

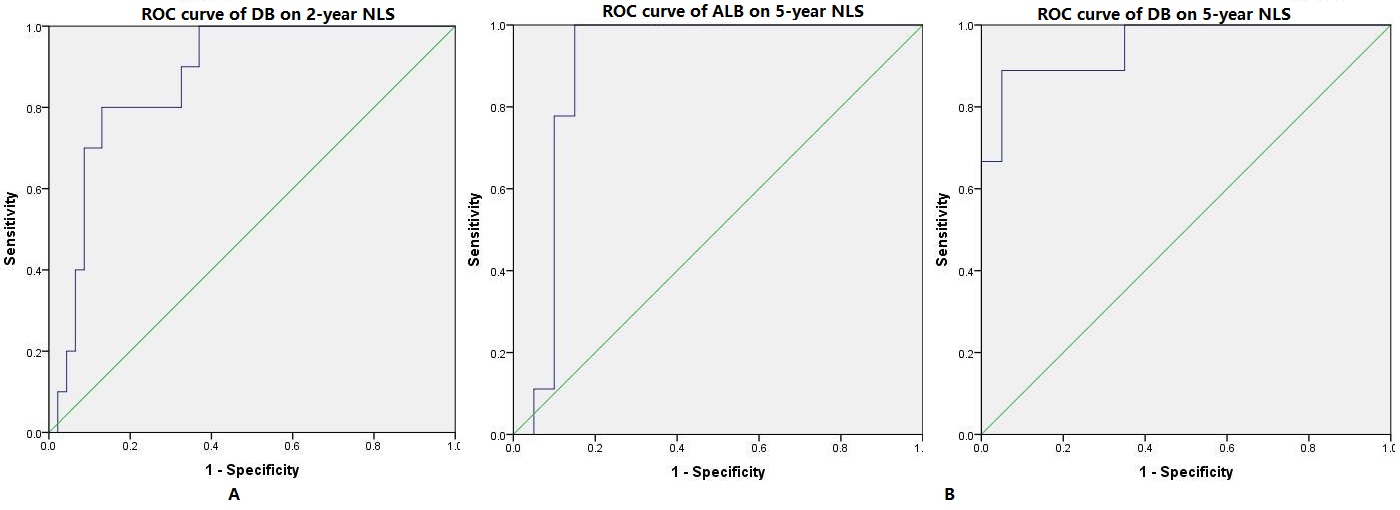

Figure S3. ROC analysis of DB on predicting 2-year NLS (A) , and ALB and DB on 5-year NLS (B)
